# Brain Tumor Detection Using Deep Learning

**DOI:** 10.1101/2022.01.19.22269457

**Authors:** Dheiver Santos, Ewerton Santos

## Abstract

A brain tumor is understood by the scientific community as the growth of abnormal cells in the brain, some of which can lead to cancer. The traditional method to detect brain tumors is nuclear magnetic resonance (MRI). Having the MRI images, information about the uncontrolled growth of tissue in the brain is identified. In several research articles, brain tumor detection is done through the application of Machine Learning and Deep Learning algorithms. When these systems are applied to MRI images, brain tumor prediction is done very quickly and greater accuracy helps to deliver treatment to patients. These predictions also help the radiologist to make quick decisions. In the proposed work, a set of Artificial Neural Networks (ANN) are applied in the detection of the presence of brain tumor, and its performance is analyzed through different metrics.

## 1.0 Introduction

The brain is one of the Organs most important organs in the human body and it is responsible for our ability to Think, Voluntary Movement, Language, Judgment, and Perception. Responsible for the functions of Movement, Balance, and Posture. Without it, we would act like a ‘walking puppets’. The word cerebellum comes from the Latin for “small brain”. A brain tumor is characterized by the growth of a tumor in the brain, distinguishing it as benign (non-cancerous) or malignant (cancerous). It can originate in the brain itself or come from another part of the body, called metastasis. Secondary brain tumors, or metastases, are more common than primary brain tumors. Around half of metastases from lung or breast cancer affect about 250,000 people a year worldwide, which corresponds to less than 2% of all cancer cases. Young people under the age of 15, brain tumors are the second leading cause of cancer. For this, magnetic resonance imaging (MRI) is widely used by radiologists to analyze brain tumors.[1,2] The result of the analysis carried out in this article reveals whether the brain is normal or diseased by applying deep learning techniques.

In this article, Neural Networks are used in classifying normal and tumor brains. An artificial neural network is composed of several processing units, whose operation is quite simple. These units are usually connected by communication channels that are associated with a certain weight. Units only perform operations on their local data, which are inputs received by their connections. [2] The intelligent behavior of an Artificial Neural Network comes from the interactions between the network’s processing units. There will be an input and output layer, while there can be any number of hidden layers. In the learning process, weight and bias are added to each layer’s neurons depending on input features and previous layers (for hidden layers and output layers). [3] A model is trained based on the activation function applied to the input features and the hidden layers where more learning takes place to reach the expected output.

This article focuses on creating a neural network for classifying healthy and unhealthy systems.

## 2.0 Data set

The dataset is from the Kaggle website (public dataset - Hundreds of organizations use CC0 to dedicate their work to the public domain) (https://www.kaggle.com/jakeshbohaju/brain-tumor). This dataset contains MRI images of brain tumors made available by scientist Jakeshbohaju Jakesh Bohaju. There are two folders, where one represents the normal brain image and the other represents the tumor images. Totally there are 3.762 images in both these folders. Figure 1 shows the sample normal and brain tumor image. Totally 1683 tumorous and 2079 non-tumorous images were taken.

**Figure 1.**
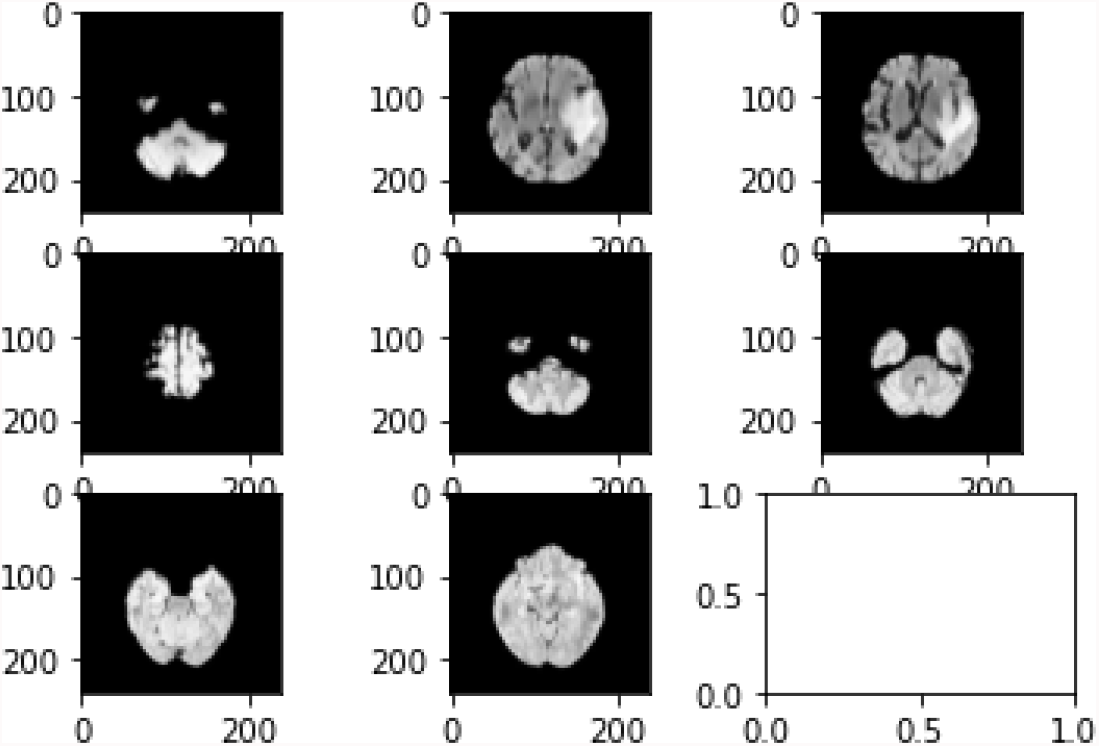
shows the sample normal and brain tumor image.

## 3.0 Deep learning

Deep Learning is one of the Artificial Intelligence technologies that has changed the market and the way machines work. Through this innovation, we have made advances in many applications in our daily lives, which shows a path to the future of our interactions with machines and services.

Deep learning, is the area of artificial intelligence related to the search for a set of rules and procedures to allow machines to act and make decisions based on data, rather than being explicitly programmed to perform a certain task.[4,5]

In this way, when analyzing a large volume of information, they can identify patterns and make decisions with the help of models. This makes machines capable of making predictions through data processing.

In this work, step by step, we use the data strategy below

1. Importing our libraries
2. Get a closer insight into our dataset
3. Get our data ready
4. Plotting some samples from the dataset
5. Preprocessing our dataset before using MobileNet
6. Splitting the dataset to the training set (80%) and testing set (20%)
7. Build model (MobileNet)

In more detail, MobileNets are a convolution class of neural networks designed by Google researchers. They were initially designed to facilitate the use of your mobile device’s resources, where part of the information is processed in the cloud, thus saving your cell phone’s computing resources.

In this work, MobileNetV2 [6] was used, which is a convolutional neural network architecture that seeks to perform well on mobile devices. It is based on an inverted residual structure where the residual connections are between the bottleneck layers. The intermediate expansion layer uses lightweight depthwise convolutions to filter features as a source of non-linearity. In this Figure 2, we detail the number of parameters that were trained (protocol).

**Figure 2.**
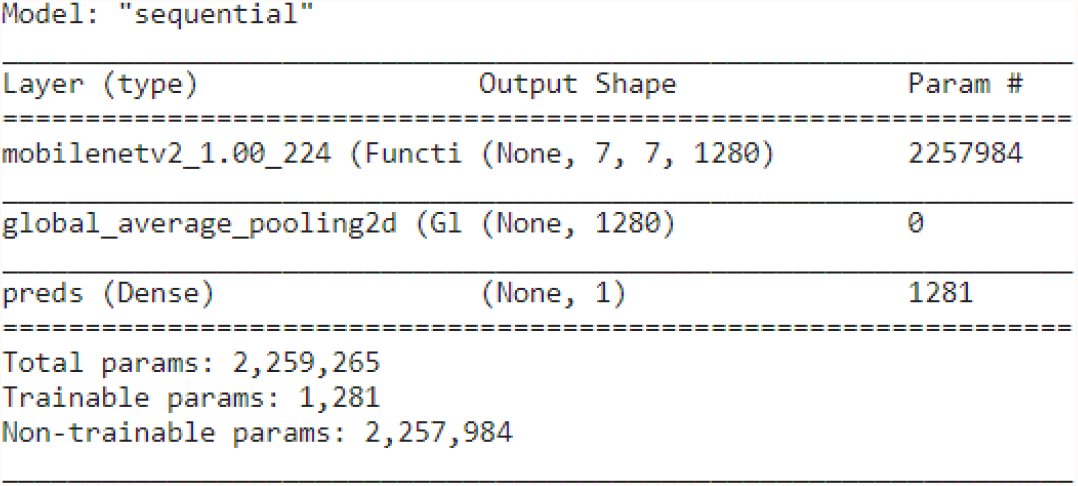
modeling strategy

## 4.0 Experimental Result Analysis

The image class labels are also generated and stored in a variable target_classes = [‘No Tumor’,’Tumor’] which is also an ndarray. The images are then added inside the dataframe. The image dataset is divided into training, validation, and testing datasets. Figure 3 represents the accuracy and loss obtained when the ANN model is applied to the training and test dataset when the ANN model is applied to the training data for fifty epochs: loss: 0.2179 - accuracy: 0.9172 - val_loss: 0.2708 - val_accuracy: 0.8924. For the validation phase we have :’precision’: 0.892, ‘recall’: 0.892, ‘f1-score’: 0.892.

**Figure 3.**
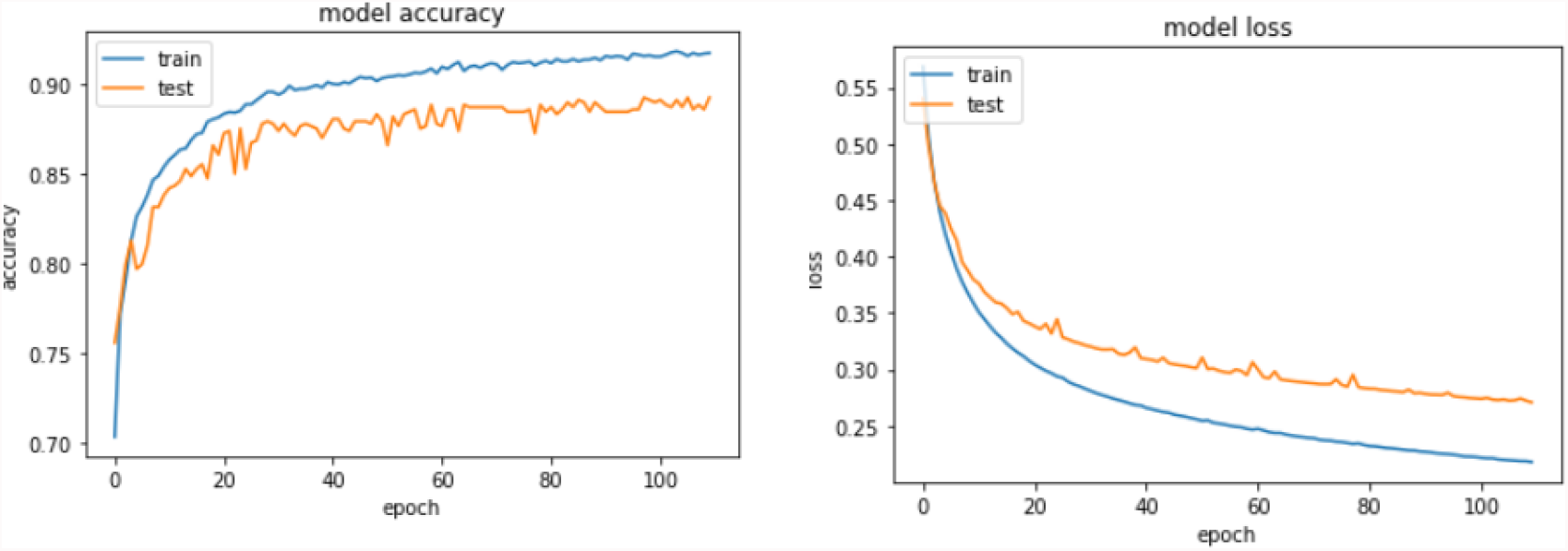
Comparing training/validation accuracy and loss of ANN model.

The model is evaluated by applying the test image dataset. The confusion matrix for the predicted output is given as in the following Figure 4. The output of making predictions for the testing and validation given below.

**Figure 4.**
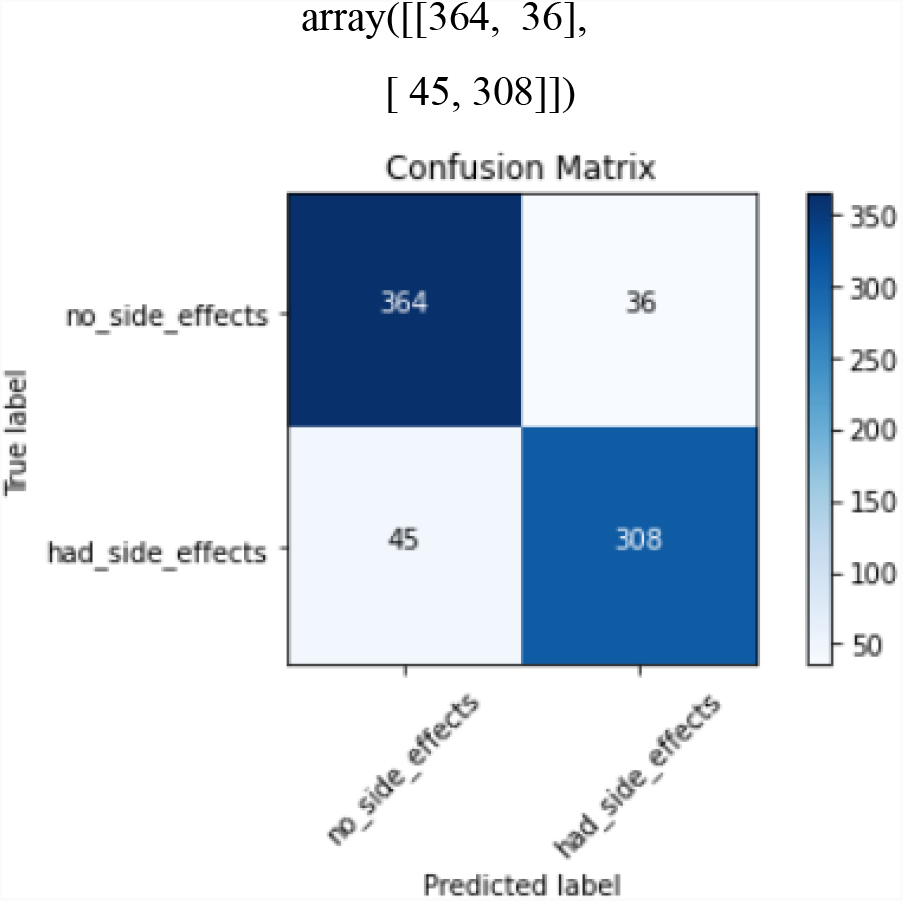
Confusion matrix for RNN model.

**Figure 5.**
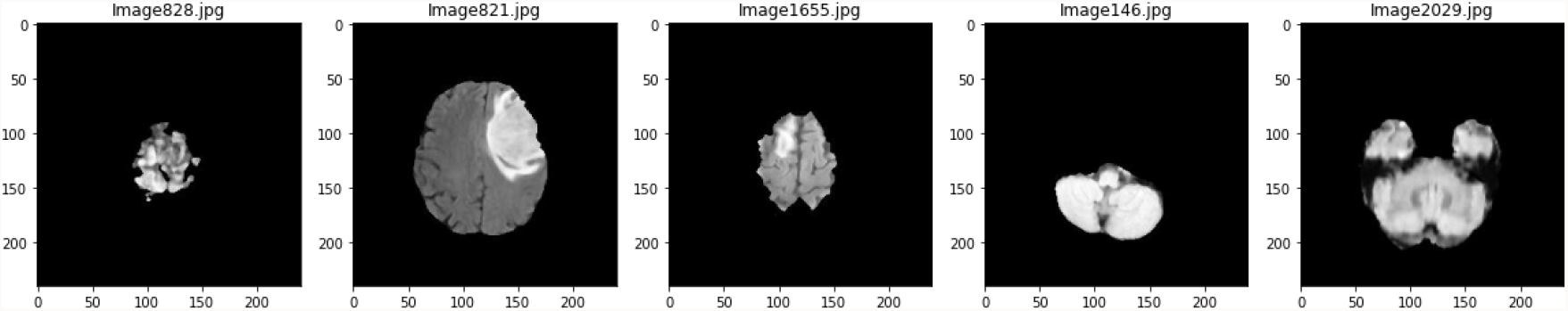
Brain Tumor Detection Using Deep Learning

In the table above, we have the comparison of the predicted value by the model with the real value of an observation. As the names suggest:

> *True Positives (TP) are observations whose actual value is positive and the predicted value is positive, i*.*e*., *the model got it right. In this work 364 images were evaluated*.
>
> *True Negatives (TN) are observations whose actual value is negative and the predicted value is negative, that is, the model got it right. In this work 308 images were evaluated*.
>
> *False Positives (FP) are cases in which the correct result is negative, however, the result obtained is positive, that is, the model was wrong. In this work 36 images were evaluated*.
>
> *False Negatives (FN) are cases in which the correct result is positive but the result obtained is negative, that is, the model was wrong. In this work 45 images were evaluated*.

## 5.0 Conclusion

Deep Learning is considered one of the best techniques in dataset analysis and artificial intelligence, despite losing a little in explainability. The Network predicts by reducing the size of the image without losing the information needed to make predictions. The model generated here yields 89% test accuracy and this can be increased by providing more image data. The same can be done by applying image enhancement techniques and analyzing the performance of other AI techniques. Soon, we will see several companies and business models proposed to help radiologists and physicians in the rapid and agile detection of brain tumors with the help of AI.

## Data Availability

All data produced in the present study are available upon reasonable request to the authors

